# Impact of COVID-19 Second Wave on Healthcare Networks in the United States

**DOI:** 10.1101/2020.07.11.20151217

**Authors:** Emad M. Hassan, Hussam Mahmoud

## Abstract

The risk of overwhelming healthcare systems from a second wave of COVID-19 is yet to be quantified. Here, we investigate the impact of different reopening scenarios of states around the U.S. on COVID-19 hospitalized cases and the risk of overwhelming the healthcare system while considering resources at the county level. We show that the second wave might involve an unprecedented impact on the healthcare system if an increasing number of the population becomes susceptible and/or if the various protective measures are discontinued. Furthermore, we explore the ability of different mitigation strategies in providing considerable relief to the healthcare system. The results can aid healthcare planners, policymakers, and state officials in making decisions on additional resources required and on when to return to normalcy.

**One Sentence Summary:** A second wave of COVID-19 will have an unprecedented impact on the healthcare system.

## Main

The COVID-19 pandemic has, to date, shown a devastating impact on our society with more than 8 million confirmed cases and total fatalities exceeding 430,000 (as of 15^th^ June, 2020) ^1^. Global efforts such as early lockdown, travel bans, among others ^2,3^ have been successful in containing the spread of the virus as evident by the reduction in the number of people infected and deceased and the subsequent relief of demand on healthcare systems ^4^. Despite their effectiveness, these strong measures led to many far-reaching and dire consequences including economic ^5,6^ and mental health crises ^7,8^. Driven by these challenges, different countries utilized various criteria and plans to ease previously applied measures and gradually return to normalcy ^9^. Nevertheless, reopening communities after pandemics comes with the risk of triggering a second pandemic wave, which might be more aggressive than the first wave ^10^ and could potentially be followed by other waves.

The concern over a second wave or spikes of the pandemic has been the subject of various national and international debates. In fact, the second wave of infected cases has already been recorded in different Asian countries including the new outbreak in Beijing and Iran while spikes are noticed in the European countries after easing their lockdown ^1^. In the U.S., the country with the highest number of confirmed COVID-19 cases ^1^ (see **Fig. 1**(**a**)), had recently experienced a sudden increase in the number of daily confirmed cases in many states during the first two weeks of June 2020. Common among these states, such as Arizona, is the fact that they had removed the stay-at-home order, causing a large jump in the confirmed daily cases (see **Fig. 1**(**b**)). A spike in the daily confirmed cases, resulting from lifting most of the stay-at-home orders, can be observed in more than 13% of the U.S. counties as shown in **Fig. 1**(**c**). These spikes could be indicators of second waves, requiring more protective measures and in some cases a return to complete lockdown. Understanding the different possibilities and consequences of early easing or removing the previously imposed protective measures is critical for devising effective planning and mitigation policies for reducing social and economic consequences.

**Fig 1.**
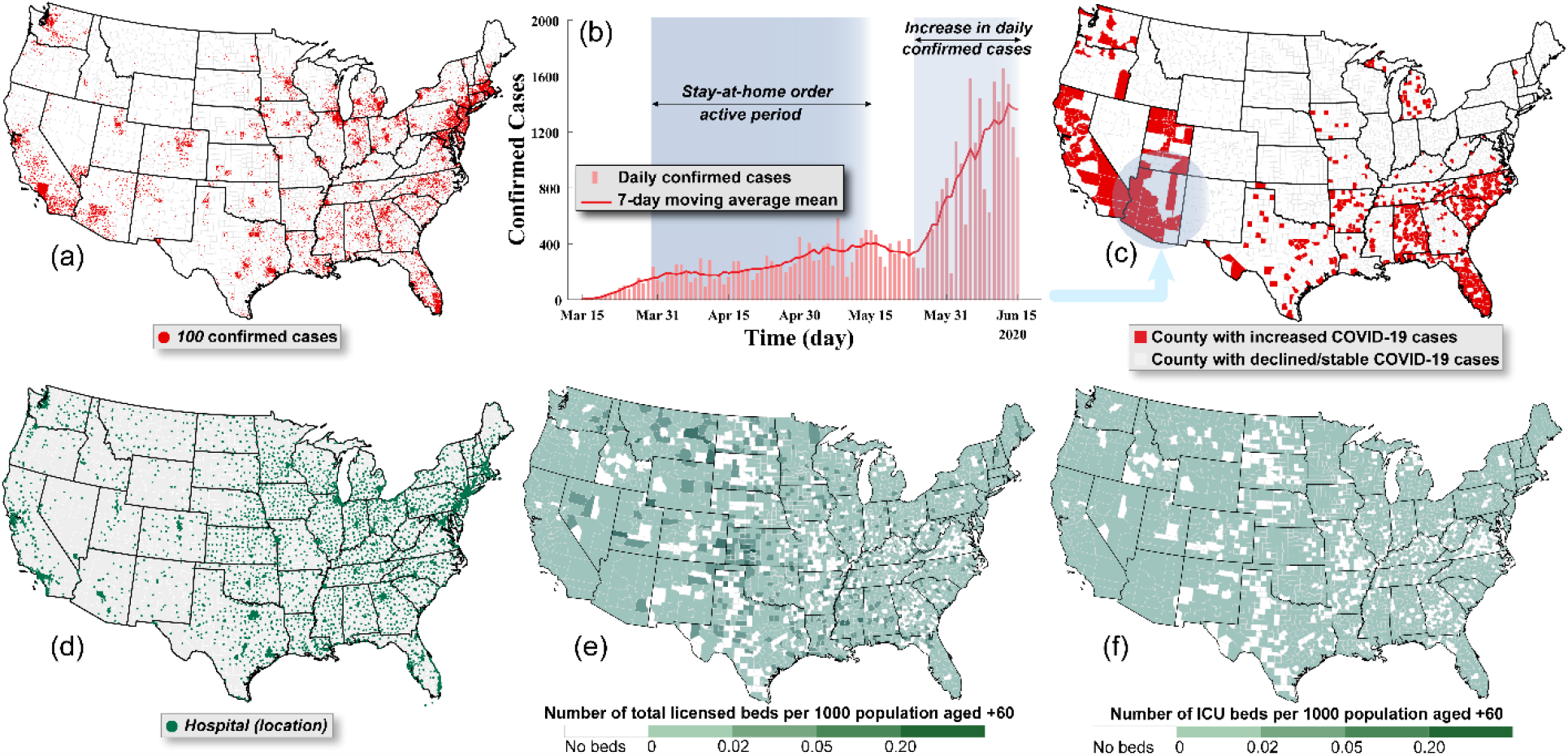
**a**) Distribution of the total number of COVID-19 confirmed cases as of 15^th^ June, 2020 in the U.S. ^20^, **b**) the daily number of confirmed cases in Arizona with the moving seven-day average means from the initial outbreak to 15^th^ June, 2020 ^20^, **c**) the U.S. counties recording the highest daily number of confirmed cases in the first two weeks of June 2020 (we excluded the counties with mean daily cases less than 5), **d**) location of U.S. hospitals with licensed staffed beds that can be used to treat COVID-19 patients ^16^, **e**) number of licensed beds per 1000 population aged +60, and **f**) number of ICU beds per 1000 population aged +60.

The healthcare systems, among other emergency services, are the frontline in the fight against the COVID-19 pandemic. However, since the start of the pandemic, healthcare facilities have experienced tremendous strain brought by the demand exceeding their capacity, forcing hospitals to make hard choices between those who can and cannot receive treatments ^11,12^. The hospitalization services needed for COVID-19 cases are based on case criticality and can be classified into inpatient admission, intensive care unit, and ventilator machine while the hospitalization services and length of stay can be a function of the patient age ^13–15^. Failing to provide adequate and appropriate hospitalization services to those infected can increase the fatality rates, especially for critical cases. In the U.S., the total number of hospitals is 6,622 and includes about 966,564 beds, 92,777 intensive care unit (ICU) beds ^16^, and 62,000 fully-featured mechanical ventilators ^17^ (see **Fig. 1**(**d**)). The number of unoccupied beds per county, which can be calculated using the total number of beds and utilization rates for these beds ^16^ (see **Fig. S1** in the Supplementary Material (SI)) shows disparities in the distribution of the healthcare system where many U.S. counties, have no beds for any patients including COVID-19-related patients ^18^ (see Material and Methods). This is simply because no hospitals exist in these counties. These disparities can have a devastating impact on the healthcare system outcomes ^19^ especially for vulnerable populations (aged +60) as shown in **Fig. S2**, who have no access to neither inpatient nor ICU beds in their counties as shown in **Fig. 1**(**e**) and (**f**).

In this research, we investigate the possibility of a second COVID-19 wave and its expected impact on the healthcare system in the U.S. after states reopening. We perform a disease transmission analysis at the county level to estimate the expected number of hospitalization cases. We further compare the different available hospitalization resources to the estimated number of hospitalization cases to highlight the impact of a second wave on patient’s accessibility to medical services. We test different state reopening scenarios including various percentages of susceptible cases and protection rates and assess the impact of each scenario on the expected number of cases needing hospitalization service and on the number of counties that might face a surge in patients beyond their beds capacity. We also investigate the effectiveness of various mitigation strategies, including postponing states reopening date and increasing hospital capacity, on enhancing the ability of hospitals to provide services for the infected patients. Furthermore, we provide an estimate of the required numbers of hospital inpatient and ICU beds as well as mechanical ventilators in case of states reopening.

## Results

This section discusses the impact of a first COVID-19 wave on the hospital system in the U.S. and the expected impact of a probable second wave on the hospitals, which are already strained from the first wave. With a total of 2,185,699 confirmed cases, the U.S. has the highest number of active cases (See Material and Methods for the definition of active cases, ***A***) in the world as of 15^th^ June, 2020 ^1^. This high number of cases is expected to further increase during the coming weeks, as shown in **Fig. 2**(**a**), assuming the current protection rates and measures will be continued as is. This disease spread prediction, based on a modified disease transmission model (see Material and Methods), is calibrated to data collected until 15^th^ June, 2020, and is referred to as the “basic case”. **Fig. 2**(**a**) shows the prediction for the aggregated active cases, ***A***, in each county, compared with two different datasets ^1,21^. Assuming no easing or relaxing of current measures, the peak of the active cases is expected to take place in early July, reaching 1.36 million as shown in **Fig. 2**(**a**). Considering the uncertainty associated with the ratio patients from the active cases requiring hospitalization service, the expected number of inpatient and ICU admissions as well as patients requiring mechanical ventilators will be as shown in **Fig. 2**(**b**). This expected demand on the healthcare system is compared with the number of unoccupied staffed beds capacity in the U.S. This analysis shows that the capacity of healthcare system the U.S. can handle the maximum demand from COVID-19 cases resulting from the basic case scenario.

**Fig 2.**
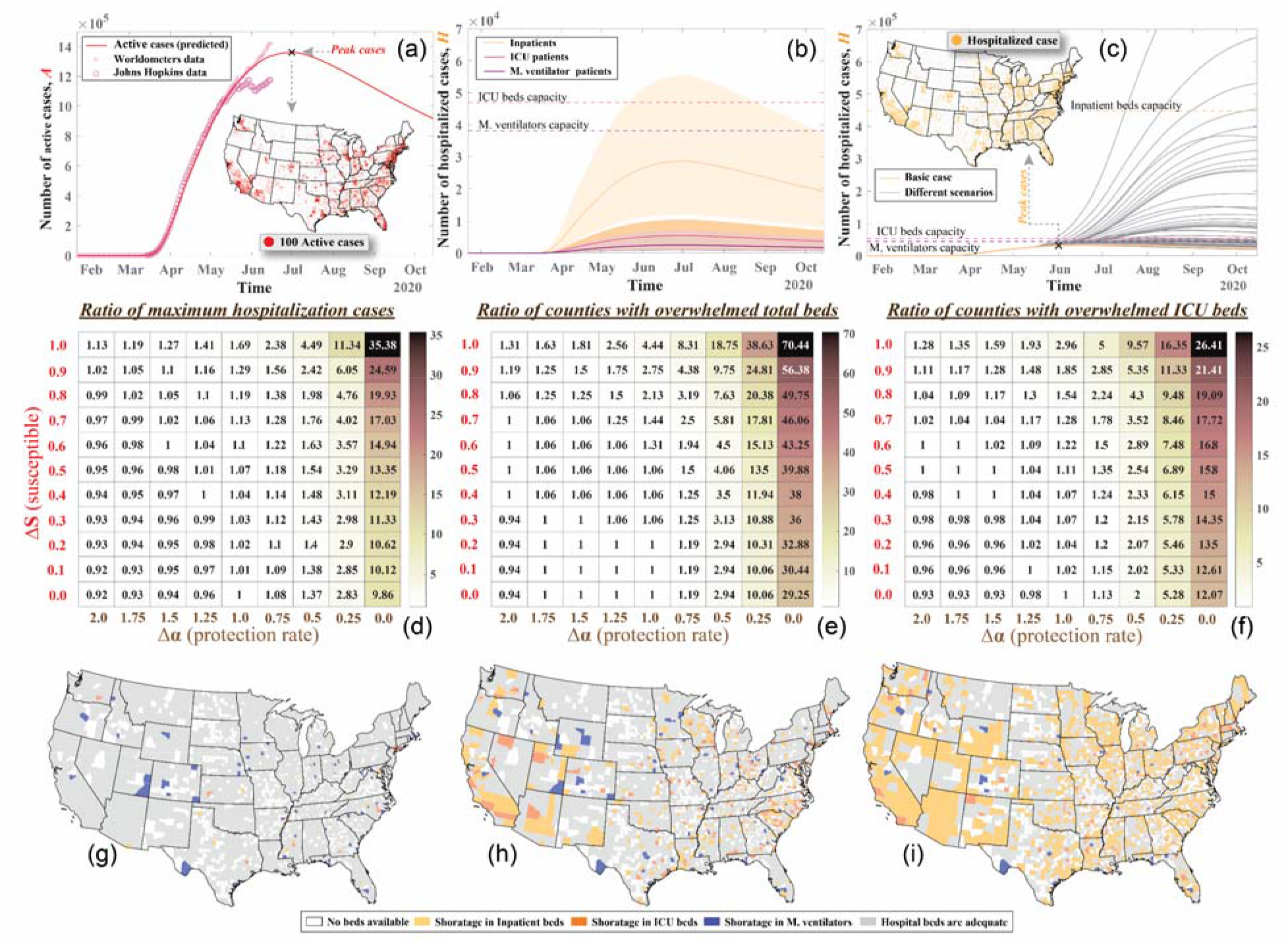
**a)** Fitting of the first wave of the active COVID-19 cases in the U.S. compared with the data collected from Worldometers ^1^ and Johns Hopkins published data ^21^ sets as well as the distribution of the maximum number of active cases, **b**) expected number of the hospitalized cases including inpatients, ICU admitted patients, and patients requiring mechanical ventilators compared with the available capacity of the U.S. beds in first two categories (the shaded area is the envelope for the 2.5 and 97.5 percentiles for each bed category). **c**) number of hospitalized cases for different state reopening scenarios compared with the basic case as well as the distribution of hospitalization cases during the peak of the basic case. The impact of different state reopening scenarios on the ratio of **d**) maximum number of cases needing hospitalization in the U.S., **e**) total number of counties with expected hospital demand exceeding the capacity, and **f**) total number of counties with expected ICU demand exceeding the capacity. Distribution of counties expected to be overwhelmed with COVID-19 patients for the **g**) basic case, **h**) fully susceptible population with 50% reduction in protection rate, and **i**) fully susceptible population with no protection.

In addition to the basic case, we investigate the impact of eliminating the protective measures on increasing the spread of the disease. Eliminating the protective measures includes deactivating the stay-at-home order, reopening schools, workplaces, and other places, and restoring the mobility rates. In this analysis, we use a generalized *SEIR* model in which we adjust the percentage of susceptible cases (***S***) and protection rate (*α*) with the time to simulate the increase in mobility and release of protective measures after the deactivation date of stay-at-home orders at each state (see **Table S1**), respectively (see Material and Methods). The percent increase in ***S*** defines the percentage of population returning to their normal daily routine. An increased *α* (Δ*α* > 1) represents more restrictions while a reduced *α* (Δ*α* < 1) indicates less restrictive measures such as easing social distancing and not imposing face masks as well as delaying the next stay-at-home order and states lockdown. The case of *α* equals zero denotes no restrictions will be applied. The results show a significant increase in hospitalized cases due to the elimination of protective measures compared with the basic case (see **Fig. 2**(**c**)). The ratios between the peak of the cases need hospitalization during each scenario and the basic case is indicated in **Fig. 2**(**d**). The figure shows that enhancing the protective measures (increasing *α*) can reduce the number of hospitalization up to 8% but releasing these measures while opening the states can be catastrophic and hospitalization can be doubled up to 35 times, which increases the demand in many counties beyond their healthcare system capacity as shown in **Fig. 2**(**e**) and (**f**). It can also be noticed from these figures that the ratio of change in protection measures (Δ*α*) is more significant on the spread of the disease than the ratio of the population returning to the normalcy (Δ***S***). In addition, releasing all the protective measures (Δ*α*=0) can increase the number of hospitalized cases ten times even without increasing the number of susceptible cases (***S***). Therefore, maintaining the protective measures is critical in reducing the number of cases and preventing overwhelming of hospitals. More details about the expected number of cases needing inpatient admission, ICU admission, and mechanical ventilators compared with available beds, in each state, can be found in **Fig. S3, Fig. S4**, and **Fig. S5** for the three scenarios discussed in the following section.

We also identify counties in the U.S. with an expected healthcare system demand exceeding the county’s staffed bed capacity as shown in **Fig. 2**(**g**), (**h**), and (**i**) for the basic case, full susceptible population and 50% protection rate, and fully susceptible population and no protection, respectively. In addition to the counties with no staffed beds for any of the three considered bed types (754 counties), the number of counties that might experience a shortage in inpatient beds is 16 {*2*.*5p*^*th*^ = 60, *97*.*5p*^*th*^ = 4}, the ICU beds is 46 {*2*.*5p*^*th*^ = 63, *97*.*5p*^*th*^ = 4}, and the mechanical ventilators is 76 {*2*.*5p*^*th*^ = 205, *97*.*5p*^*th*^ = 34} for the basic case. Most of these counties are rural and the number of patients overflow is minimal and therefore can be accommodated by patient transfer to other hospitals within each state. However, for the full susceptible population with 50% reduction in the protection rate of the basic case, the numbers of overwhelmed counties will increase to 292 {*2*.*5p*^*th*^ = 468, *97*.*5p*^*th*^ = 126} for inpatient beds, 440 {*2*.*5p*^*th*^ = 652, *97*.*5p*^*th*^ = 86} for ICU beds, and 392 {*2*.*5p*^*th*^ = 702, *97*.*5p*^*th*^ = 126} for mechanical ventilators. For the fully susceptible population and no protection, the number of overwhelmed counties will be 1,118 {*2*.*5p*^*th*^ = 1236, *97*.*5p*^*th*^ = 899} for inpatient beds, 1215 {*2*.*5p*^*th*^ = 1306, *97*.*5p*^*th*^ = 836} for ICU beds, and 1151 {*2*.*5p*^*th*^ = 1296, *97*.*5p*^*th*^ = 764} for mechanical ventilators. In these two scenarios, we show that even urban counties with a large number of staffed beds might also be overwhelmed. The distribution for counties expected to be overwhelmed with COVID-19 patients during peak cases, based on different susceptible cases and protection rates, is shown in **Fig. S6** for the scenarios summarized in **Fig. 2**(**d**).

## Discussion

In this section, we investigate different strategies that might help reduce the consequences associated with partial elimination of protective measures after state reopening. These strategies are commonly used by communities and healthcare system planners to decrease the number of hospitalized cases and/or to enhance the healthcare system during the pandemic. Here, we use the case of a fully susceptible and 50% reduction in protection rate, which we discussed earlier in the results section. The early reopening of states before controlling the disease spread and significantly reducing the number of infectious cases can have devastating impacts including a sudden increase in the number of infected cases, and in some cases might cause severe second waves of disease spread. Our analysis shows that delaying the states’ reopening process and maintaining strong mitigation measures can efficiently reduce the number of infected and hospitalized cases as well as decrease the risk of overwhelming the healthcare system with patients as shown in **Fig. 3**(**a**). While early reopening will ultimately lead to a second wave with a magnitude of 4.5 {*2*.*5p*^*th*^ = 8.7, *97*.*5p*^*th*^ = 1.7} times the basic case, appropriate timing of the reopening can result in minor waves with substantial reducing in the peak in hospitalized cases, which can crucially prevent overwhelming of the healthcare system. It can also be noticed from the analysis that the impact of reopening is a function of the number of infectious cases in the population at the reopening stage. However, this is not always the case and if no further protective measures are applied (*α*=0), this impact will be minimal.

**Fig 3.**
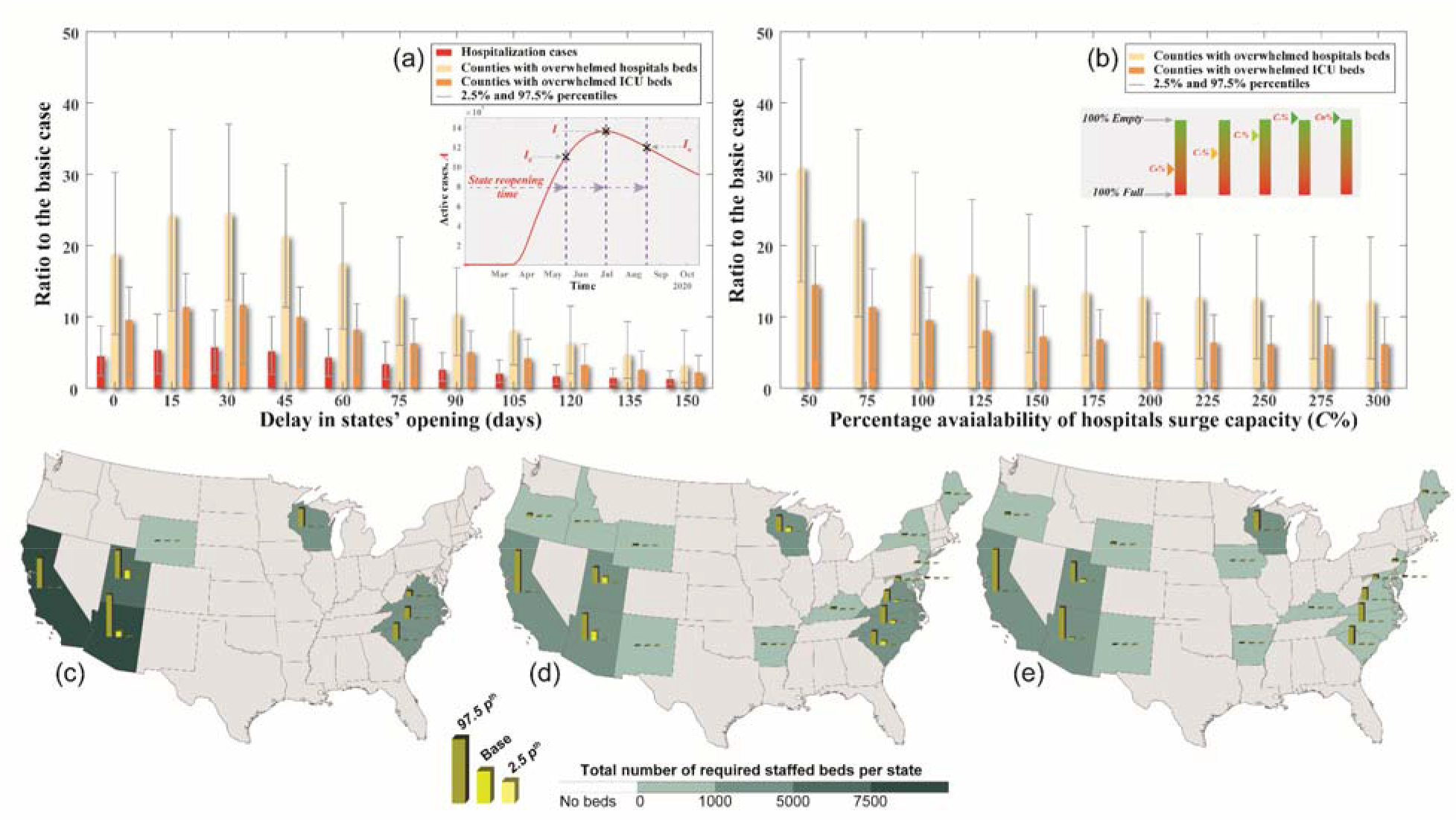
**a**) Impact of states reopening time on the expected distribution of the hospitalized cases, number of overwhelmed counties based on the total number of beds and ICU beds **b**) effect of the available surge capacity at each state on the expected number of overwhelmed counties based on the total number of beds and ICU beds. Required number of additional staffed beds per state to avoid overwhelming the healthcare system based on **c**) inpatient beds, **d**) ICU beds, and **e**) mechanical ventilators. The bar graphs represent the base, 97.5, and 2.5 percentiles while the color fill in the maps denotes the 97.5 percentile.

One of the main approaches the healthcare systems are using to manage the sudden increase in patients is to provide surge capacity ^22^. This surge in capacity can be realized by reducing the hospitalization rates of regular patients to allow more COVID patients to be admitted. The number of regular patients in each county has been documented since the beginning of the COVID-19 pandemics ^23^. In this analysis, we assume no increase in staffed beds in each hospital, but we consider more beds can be assigned for COVID-19 related patients. This approach can considerably reduce the number of overwhelmed counties as shown in **Fig. 3**(**b**); however, providing surge capacity is limited to the number of beds in each facility. When compared to the basic case, providing surge capacity can reduce the ratio of counties with overwhelmed hospitals to 12.2 {*2*.*5p*^*th*^ = 21.2, *97*.*5p*^*th*^ = 10.9} and counties with overwhelmed ICU beds to 6.1 {*2*.*5p*^*th*^ = 10.0, *97*.*5p*^*th*^ = 0.9}. On the other hand, the number of overwhelmed counties can significantly increase if the surge in capacity is limited. In such case, the ratio of counties with overwhelmed hospitals can increase to 30.9 {*2*.*5p*^*th*^ = 46.1, *97*.*5p*^*th*^ = 14.9} and counties with overwhelmed ICU beds to 14.5 {*2*.*5p*^*th*^ = 20.0, *97*.*5p*^*th*^ = 4.1} compared with the basic case.

Another method to increase the capacity of the healthcare system is to add additional staffed beds. These additional staffed beds can be added as field hospitals ^24^ or backup beds at the existing hospitals ^22^. We identify in this analysis the states that will need additional beds and quantify the number of beds that will be required. To quantify the optimal number of staffed beds needed at each US state, we first evaluate the expected maximum number of cases need hospitalization per state and assume that the healthcare system in each state can manage to treat patients from overwhelmed counties ^25^; however, in many cases, the capacity of state’s healthcare system can also be overwhelmed (see **Fig. S4**) then additional support will be required to bridge the gap between the demand and capacity of the staffed beds. **Fig. 3**(**c**), (**d**), and (**e**) show the number of staffed beds needed per state based on the basic, 2.5, and 97.5 percentiles (see material and methods) for the inpatient beds, ICUs, and mechanical ventilators, respectively. The analysis shows that the states located on the West and East Coasts are more vulnerable to hospital overwhelming and will need additional beds and mechanical ventilators if states are fully reopened and 50% reduction in protection rate take place. The required additional staffed beds and mechanical ventilators per state for other different scenarios are shown in **Fig. S7**.

In conclusion, we explored the different possibilities of the COVID-19 second wave in the U.S. and their impact on the healthcare system around the country. We used a generalized *SEIR* model to predict the number of hospitalized cases for each county considering various state reopening scenarios including partial or fully reopening while taking into account different levels of population protection rates. We identified the counties that might experience overwhelming patients demand that exceeds their healthcare system capacity. We further investigated the impact of different mitigation strategies on the number of cases that need hospitalization and the availability of the healthcare system and estimated the number of staffed beds that will be needed to overcome the expected shortage of the staffed beds.

The presented analysis concentrates on the impact of COVID-19 second wave on the healthcare system with a focus on estimating the cases that need hospitalization while considering the available resources in each county. We assumed the number of recovered cases in each county based on the recovery rates of the U.S. due to the current limitation of these data. We evaluated the uncertainty in the hospitalized cases and fitted the disease transmission model to published data to estimate the disease transmission parameters. However, utilizing more data could lower the level of uncertainties in these estimates. We assumed that the population per county is constant and we neglected the impact of the relocation between states on disease spread. Furthermore, we used published data to estimate the number of staffed beds per county and the utilization of these beds and we are not accounting for the additional staffed beds and field hospitals built after the pandemic outbreak in the U.S.

## Material and methods

### Disease transmission model

Among many other disease transmission models ^26^, the *SEIR* models are commonly used to model the COVID-19 pandemic ^27,28^. We developed a modified version of the generalized six-states *SEIR* disease transmission model ^29^ to include ten different states as follow {***S, P, E, I, Q, T, C*, V, *R, D***}_*t*_ to represent the susceptible, insusceptible, exposed, infective, self-quarantined, inpatient admitted, *ICU* admitted, cases on a mechanical ventilator, recovered, and deceased cases, respectively as a function of time, *t*. The additional four cases (***Q, T, C***, and ***V***) represent different types of confirmed cases based on their hospitalization services needs in which ***Q*** is for cases with mild or no symptoms, ***T*** is for cases needing hospital admission, ***C*** is for cases needing ICU, and ***V*** is for cases needing mechanical ventilators. These four states can be aggregated to represent the total number of active cases, ***A***, that represents all the positive (confirmed) cases with no outcomes (recovered or deceased) yet. ***T, C***, and ***V*** cases together form the total COVID-19 demand on the healthcare system, ***H***. The following model is constructed for population, *N*, of each county, *i*, in the U.S. The differential equations below are used to determine the total number in each state in county *i*.

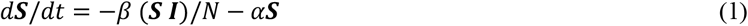

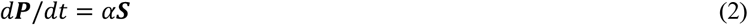

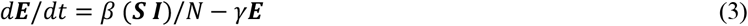

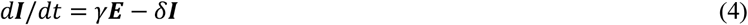

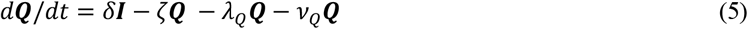

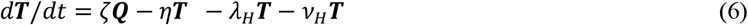

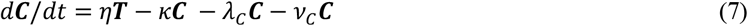

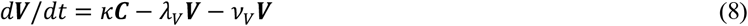

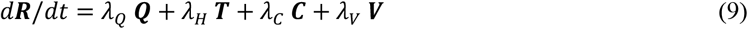

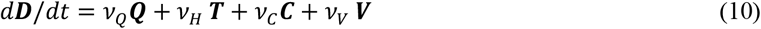

Where, *β* is the infection rate, *α* is the protection rate, *1/γ* is the average incubation period, *1/δ* is the average quarantine time, *ζ* is the hospitalization rate, *η* is the *ICU* rate, and *κ* is the mechanical ventilator rate. In addition, *λ*_*Q*_, *λ*_*H*_, *λ*_*C*_, and *λ*_*V*_ are the recovery rates for self-quarantined, inpatient, *ICU*, and mechanical ventilator cases, respectively. Moreover, *ν*_*Q*_, *ν*_*H*_, *ν*_*C*_, and *ν*_*V*_ are the death rate for the self-quarantined, inpatients, and patients in the *ICU*, and those on a mechanical ventilator, respectively. Furthermore, the basic reproduction number, *R*_*0*_, is the average number of secondary infective cases produced by one infective case in the same county during the infectious period of this case and equals to *β/δ*(*1-α*)^*t*^. In the utilized model we assume a constant population for each investigated county over the epidemic time, *N*, which satisfies the equilibrium of *N*=***S***+***P***+***E***+***I***+***Q***+***T***+***C***+***V***+***R***+***D*** at any time *t*.

Using the modified *SEIR* model and including self-quarantined and hospitalized states while accounting for the impact of protective measures allows for more reliable fitting and forecasting of the COVID-19 disease spread. Assigning positive values to the protection rate, *α*, is utilized to simulate different protective measures including lockdown, social distancing, wearing protective masks, etc. To simulate state reopening, we model different percentages of the population who return to normalcy and become non-protected population (Δ***S***) in the reopened counties by increasing the number of susceptible populations at the stay-at-home deactivation time (see **Table S1**). While modeling the strengthening or easing of the protective measures in each county is realized by changing the protection rate as a ratio of the protection rate for the basic case (Δ*α*) at the county reopening date (see **Table S1**). Shifting the *α* can change the rate of disease spread and the basic reproduction number, *R*_*0*_, which can be reduced with time ^29^ using protective measures^30^

Estimation of the model parameters (*β, α, γ, δ*, and *ν*(s)) is made by fitting the published data for confirmed and deceased ^20^, while λ(s) are estimated as a time-dependent parameter from the published US recovery data ^1,21^ and assumed to be similar for all the US counties. Initial values for the parameter estimation are assumed based on previous studies ^14,31^ and CDC reports ^13,30^. During the state’s reopening, *α* is modified based on the investigated scenario. We simulated the *ζ* as gamma distribution with 0.025, 6.33, and 0.004 for base, shape, and scale parameters, respectively, while for *η* we used gamma distribution with 0.16, 6.13, and 0.02 for base, shape, and scale parameters, respectively, and for *κ* we utilized beta distribution with 0.46, 5.22, and 3.08 for base, shape, and scale parameters, respectively ^14,31^. These distributions are used to model the uncertainty associated with the number of different hospitalization cases, in which we use Monto-Carlo simulations with 100,000 trials.

### Healthcare system model

Modeling the capacity of the healthcare system in the US is based on the published data for all the US hospitals including hospital location, the number of staffed beds and ICU beds, and the utilization ratio of these beds. We aggregate these data to calculate the number of total inpatient and ICU beds in each county, while, due to the limited data about the number of mechanical ventilators per county, we assume that the number of mechanical ventilators per the ICU bed is constant and estimated based on the recently published estimates of ventilators in the US ^17^. To simulate the available surge capacity (unoccupied beds) per county, we use the number of staffed beds while considering the potential increase in these beds and multiply it by the utilization rates. The capacity calculations are then verified with the data from the CDC dashboard ^32^. These beds are used in our analysis for COVID-19 cases require hospitalization service.

COVID-19 patients from counties with no staffed beds are redistributed to hospitals within the same state that has unoccupied beds. If no beds are available in the whole state, the patients from these counties are added to the total cases at the county where their most probable hospital is located using the framework presented by Hassan and Mahmoud ^33^ with assumptions to overcome the data limitation related to the hospital functionality that is not simulated in this study.

## Data Availability

All data used in the analysis are included in the paper and the Supplementary Materials. Additional data related to this paper may be requested from the corresponding author. The disease transmission model components have been described in the paper and the Supplementary Materials and will be made available from the corresponding author on request.

## Acknowledgment

Funding for this study was in part provided by the cooperative agreement 70NANB15H044 between the National Institute of Standards and Technology (*NIST*) and Colorado State University. The content expressed in this paper are the views of the authors and do not necessarily represent the opinions or views of *NIST* or the U.S Department of Commerce.

## Contributions

Emad M. Hassan and Hussam Mahmoud conceived the idea and contributed to the final version of the manuscript. Emad M. Hassan carried out the simulations. Hussam Mahmoud supervised the project.

## List of Supplementary Materials

**Fig. S1**.

**Fig. S2**.

**Table S1**.

**Fig. S3**.

**Fig. S4**.

**Fig. S5**.

**Fig. S6**.

**Fig. S7**.

## Supplementary Materials

Attached file.

## References

1. Worldometers. COVID-19 Coronavirus pandemic. https://www.worldometers.info/coronavirus/ (2020).

2. Hsiang, S. et al. The effect of large-scale anti-contagion policies on the COVID-19 pandemic. Nature (2020) doi:10.1038/s41586-020-2404-8.

3. Matrajt, L. & Leung, T. Evaluating the effectiveness of social distancing interventions against COVID-19. Emerg. Infect. Dis. 8, (2020).

4. Branas, C. C. et al. Flattening the curve before it flattens us: hospital critical care capacity limits and mortality from novel coronavirus (SARS-CoV2) cases in US counties. medRxiv 2020.04.01.20049759 (2020) doi:10.1101/2020.04.01.20049759.

5. Guan, D. et al. Global supply-chain effects of COVID-19 control measures. Nat. Hum. Behav. (2020) doi:10.1038/s41562-020-0896-8.

6. Gopinath, G. The Great Lockdown: Worst Economic Downturn Since the Great Depression. https://blogs.imf.org/2020/04/14/the-great-lockdown-worst-economic-downturn-since-the-great-depression/ (2020).

7. Panchal, N. et al. The Implications of COVID-19 for Mental Health and Substance Use. https://www.kff.org/coronavirus-covid-19/issue-brief/the-implications-of-covid-19-for-mental-health-and-substance-use/ (2020).

8. Pereira-Sanchez, V. et al. COVID-19 effect on mental health: patients and workforce. The lancet. Psychiatry 7, e29–e30 (2020).

9. Gilbert, M., Dewatripont, M., Muraille, E., Platteau, J.-P. & Goldman, M. Preparing for a responsible lockdown exit strategy. Nat. Med. 26, 640–642 (2020).

10. Taubenberger, J. K. & Morens, D. M. 1918 Influenza: the Mother of All Pandemics. Emerg. Infect. Dis. 12, 15–22 (2006).

11. Rosenbaum, L. Facing Covid-19 in Italy — Ethics, Logistics, and Therapeutics on the Epidemic’s Front Line. N. Engl. J. Med. 1969–73 (2020) doi:10.1056/NEJMp2009027.

12. Zhang, Z., Yao, W., Wang, Y., Long, C. & Fu, X. Wuhan and Hubei COVID-19 mortality analysis reveals the critical role of timely supply of medical resources. J. Infect. 81, 147–178 (2020).

13. CDC COVID-19 Response Team. Severe Outcomes Among Patients with Coronavirus Disease 2019 (COVID-19) - United States, February 12-March 16, 2020. MMWR. Morbidity and mortality weekly report vol. 69 (2020).

14. Zhou, F. et al. Clinical course and risk factors for mortality of adult inpatients with COVID-19 in Wuhan, China: a retrospective cohort study. Lancet 395, 1054–1062 (2020).

15. Lauer, S. A. et al. The Incubation Period of Coronavirus Disease 2019 (COVID-19) From Publicly Reported Confirmed Cases: Estimation and Application. Ann. Intern. Med. 2019, (2020).

16. Definitive Healthcare. USA Hospital Beds. https://coronavirus-resources.esri.com/datasets/definitivehc::definitive-healthcare-usa-hospital-beds?geometry=97.382%2C-16.820%2C-122.344%2C72.123&selectedAttribute=NUM_ICU_BEDS (2020).

17. Society of Critical Care Medicine. Shortage of ICU Providers Who Operate Ventilators Would Severely Limit Care During COVID -19 Outbreak. (2020).

18. Miller, I. F., Becker, A. D., Grenfell, B. T. & Metcalf, C. J. E. Disease and healthcare burden of COVID-19 in the United States. Nat. Med. (2020) doi:10.1038/s41591-020-0952-y.

19. Committee on Guidance for Designing A National Healthcare Disparities Report. Guidance for the National Healthcare Disparities Report. Guidance for the National Healthcare Disparities Report (National Academy of Sciences, 2002). doi:10.17226/10512.

20. USAFACTS. Coronavirus Locations: COVID-19 Map by County and State. https://usafacts.org/visualizations/coronavirus-covid-19-spread-map/ (2020).

21. Dong, E., Du, H. & Gardner, L. An interactive web-based dashboard to track COVID-19 in real time. Lancet Infect. Dis. 3099, 19–20 (2020).

22. Carenzo, L. et al. Hospital surge capacity in a tertiary emergency referral centre during the COVID-19 outbreak in Italy. Anaesthesia 75, 928–934 (2020).

23. Hartnett, K. P. et al. Impact of the COVID-19 Pandemic on Emergency Department Visits — United States, January 1, 2019–May 30, 2020. Morbidity and Mortality Weekly Report vol. 69 (2020).

24. Chen, S. et al. Fangcang shelter hospitals: a novel concept for responding to public health emergencies. Lancet 395, 1305–1314 (2020).

25. Denver Health. National Hospital Available Beds for Emergencies and Disasters (HAvBED) System: Final Report. AHRQ Publication No. 05-0103. (2005).

26. Grassly, N. C. & Fraser, C. Mathematical models of infectious disease transmission. Nat. Rev. Microbiol. 6, 477–487 (2008).

27. Wu, J. T., Leung, K. & Leung, G. M. Nowcasting and forecasting the potential domestic and international spread of the 2019-nCoV outbreak originating in Wuhan, China: a modelling study. Lancet 395, 689–697 (2020).

28. Kucharski, A. J. et al. Early dynamics of transmission and control of COVID-19: a mathematical modelling study. Lancet Infect. Dis. 3099, 1–7 (2020).

29. Peng, L., Yang, W., Zhang, D., Zhuge, C. & Hong, L. Epidemic analysis of COVID-19 in China by dynamical modeling. 1–18 (2020).

30. CDC. Implementation of Mitigation Strategies for Communities with Local COVID-19 Transmission. www.cdc.gov/COVID19 (2020).

31. Weissman, G. E. et al. Locally Informed Simulation to Predict Hospital Capacity Needs During the COVID-19 Pandemic. Ann. Intern. Med. (2020) doi:10.7326/m20-1260.

32. Centers for Disease Control and Prevention. COVID-19 Module Data Dashboard – Patient Impact and Hospital Capacity Pathway. National Healthcare Safety Network https://www.cdc.gov/nhsn/covid19/report-patientimpact.html (2020).

33. Hassan, E. M. & Mahmoud, H. An integrated socio-technical approach for post-earthquake recovery of interdependent healthcare system. Reliab. Eng. Syst. Saf. (2020) doi:10.1016/j.ress.2020.106953.

